# What works in educational interventions for teams caring for children and young people admitted to general acute paediatric wards for mental health? A systematic review

**DOI:** 10.1101/2025.10.27.25338855

**Authors:** Olatokunbo Sanwo, Antonia Rich, Isabelle Pomfret, James Downs, Sophie Davis, James R Dearden, Mark Buchanan, Lee Omar, Ann Griffin, Lee D Hudson

## Abstract

**Objective:** To systematically review educational interventions for staff working on acute paediatric wards aimed at improving the care of children and young people (CYP) admitted for mental health support.

**Design:** We conducted a systematic search of PubMed, ERIC, PsychInfo, and Web of Science from 2000 to September 2025 using terms related to *healthcare professionals, training/education, mental health*, and *children/young people*. PRISMA guidelines were followed. Interventions were classified using the Kirkpatrick model and the Behavioural Change Technique Taxonomy (BCT v1) to identify mechanisms underpinning effective change.

**Results:** We found nine studies met inclusion criteria, across a range of teaching methodologies, and settings. Most interventions were designed and applied for nurses. Overall quality of evidence was poor. Although no single intervention could be recommended, analysis identified potentially useful components tailored to different learner needs, mapped through the Kirkpatrick and BCT frameworks. A notable gap was a lack of co-design with CYP and carers with lived experience and the absence of strategies to engage ambivalent or reluctant learners who might not attend educational interventions voluntarily.

**Conclusions:** Existing educational interventions contain elements that may support behavioural and practice change among staff caring for CYP with mental health needs. However, future research should prioritise high-quality, framework-based evaluations developed through co-design with CYP and carers. Interventions should also address how to engage learners who may be reluctant to participate in training for this area of clinical practice.

## Introduction

General, acute paediatric wards are important settings for children and young people (CYP) presenting to hospital with a deterioration in their mental health and are too unwell to be at home.(1, 2) A recent cohort study reported a 65% increase in admissions to acute paediatric wards for primary mental-health causes in England between 2012 and 2022, with increasing lengths of stay.(3) Given limited evidence for interventions to prevent or reduce such admissions to medical wards,(4, 5) such admissions will remain a significant feature of inpatient paediatric care for a foreseeable future.

The independent Healthcare Services Safety Investigation Branch (HSIB) has warned that young people with complex mental health needs are placed at significant risk when admitted to general children’s wards in the UK, citing inadequate training and skills amongst staff as key areas of risk and areas for improvement.(6) The range of presentations for such admissions are broad, requiring skill sets in supporting patients with risk of self-harm,(7) challenging behaviours,(8) and medical complications arising from starvation, as may occur amongst CYP with eating disorders.(9) The demands of complex interventions such as physical restraint(10) can also pose significant challenges for paediatric ward staff, who may lack specific mental health training and feel ill-equipped to provide safe and appropriate care,(11–14) especially as access to specialist mental health professionals is frequently limited.(15, 16) Some ward team members may feel that providing mental health care lies beyond their remit.(1, 17) Negative experiences for staff in these settings also likely contribute to burnout, reduced well-being, and workforce retention.(1) Importantly, the quality of care provided to CYP admitted with mental health concerns is also known to have a direct impact on their experience of an admission.(1, 18)

Addressing these challenges calls for enhanced education and training for paediatric staff, with coordinated actions required across local, regional and national levels. The design and implementation of educational interventions in this arena should follow evidence-based approaches, grounded in educational theory, to maximise effectiveness, minimise failure, and ensure the efficient use of resources.(19, 20) Systematic reviews of educational interventions for healthcare professionals are valuable for identifying strengths and limitations in existing literature and informing the design of future programmes.(21–24) To date, however, no published synthesis has examined educational interventions aimed at improving the ability of paediatric staff to manage mental health concerns on acute general paediatric inpatient settings. We therefore conducted a systematic review of the published literature to identify such interventions and determine which approaches, or their components, have been effective in achieving change in learners.

## Methods

We adhered to PRISMA guidelines.(25)

### Eligibility Criteria

We included peer-reviewed, published studies in English that reported on educational interventions with one or more non-specialist mental-health health professionals working in acute paediatric ward settings. Eligible studies evaluated educational interventions designed to improve the care of inpatients admitted with mental health concerns, whether focussed on general management of mental health in acute settings or on specific mental health presentations commonly seen in paediatric wards (e.g. self-harm, eating disorders and behavioural disturbances).

We excluded studies that did not describe interventions sufficiently to understand their methodology, or that lacked temporal measures of change (i.e. pre- and post-intervention). Studies involving combinations of specialist and non-specialist mental health staff were excluded if results for non-specialists could not be identified separately.

### Search method for identification of studies

We searched PubMed, The Education Resources Information Centre (ERIC), PsychInfo, and Web of Science (from 2000 to September 1^st^ 2025). Search terms were developed with a clinical librarian to capture “*healthcare professionals*”, “*training/education*”, “*mental health*”, and “*children/young people*”. Specific search terms for each database are provided in Appendix I. We also hand-searched reference lists of included papers for any additional relevant publications.

Searches at title, abstract, and full text level were conducted independently by two researchers (OS and IP). Any potential differences were resolved through discussion with a third reviewer (LDH), though no differences occurred.

### Quality Assessment

Two reviewers (OS and AR) independently assessed the quality of included studies using the Modified Medical Education Research Study Quality Intervention (MMERSQI),(26) which comprises 12 items and 43 sub-items evaluating seven domains: study design, sampling, setting, types of data, validity of evaluation instrument(s),data analysis, and outcomes(27). First round Inter-rater reliability for quality assessment was calculated as the number of disagreements divided by total number of assessments. Any remaining differences were resolved by discussion, with input from a third reviewer (LDH) where required.

### Analysis

We found insufficient studies for meta-analysis, therefore findings are presented narratively. In addition to describing the interventions and reported learner outcomes, each intervention was assessed and coded using the Behavioural Change Technique Taxonomy(BCT) version 1, a recognised framework for standardising and identifying the behavioural change components of interventions.(28) We also assessed each study using Kirkpatrick’s model, an established tool for appraising the effectiveness of educational interventions across four levels of learning.(29, 30) A visual summary of the key domains from both frameworks applied in the analysis is presented in figure 1.

**Figure 1.**
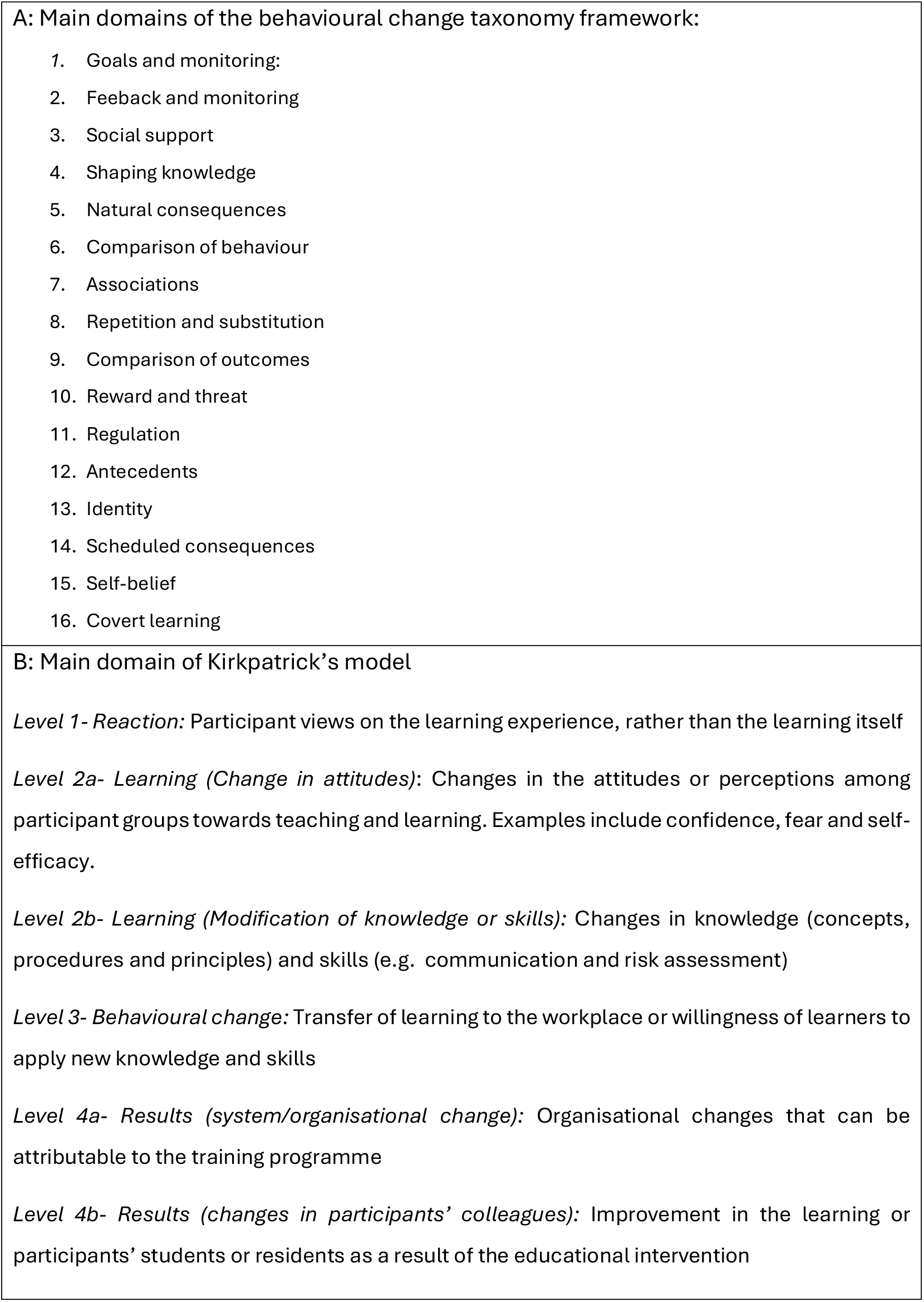
(A)Summaries of main domains of Behavioural Change Taxonomy (28) and (B) Kirkpatrick’s model of evaluation(30, 54) Full texts screened for eligibility (n = 91)

Two researchers with lived experience of being admitted to a general children’s ward with mental health concerns collaborated in assessing the relevance and interpretation of findings.

## Results

### Description of included studies

Search results are summarised in Figure 2. Nine studies met inclusion criteria (summarized in Table 2). Papers were published between 2012 and 2023, originating from America (3),(31–33) the UK (3),(34–36) Canada (1),(37) Australia (1),(38) and Nigeria (1).(39)

**Figure 2.**
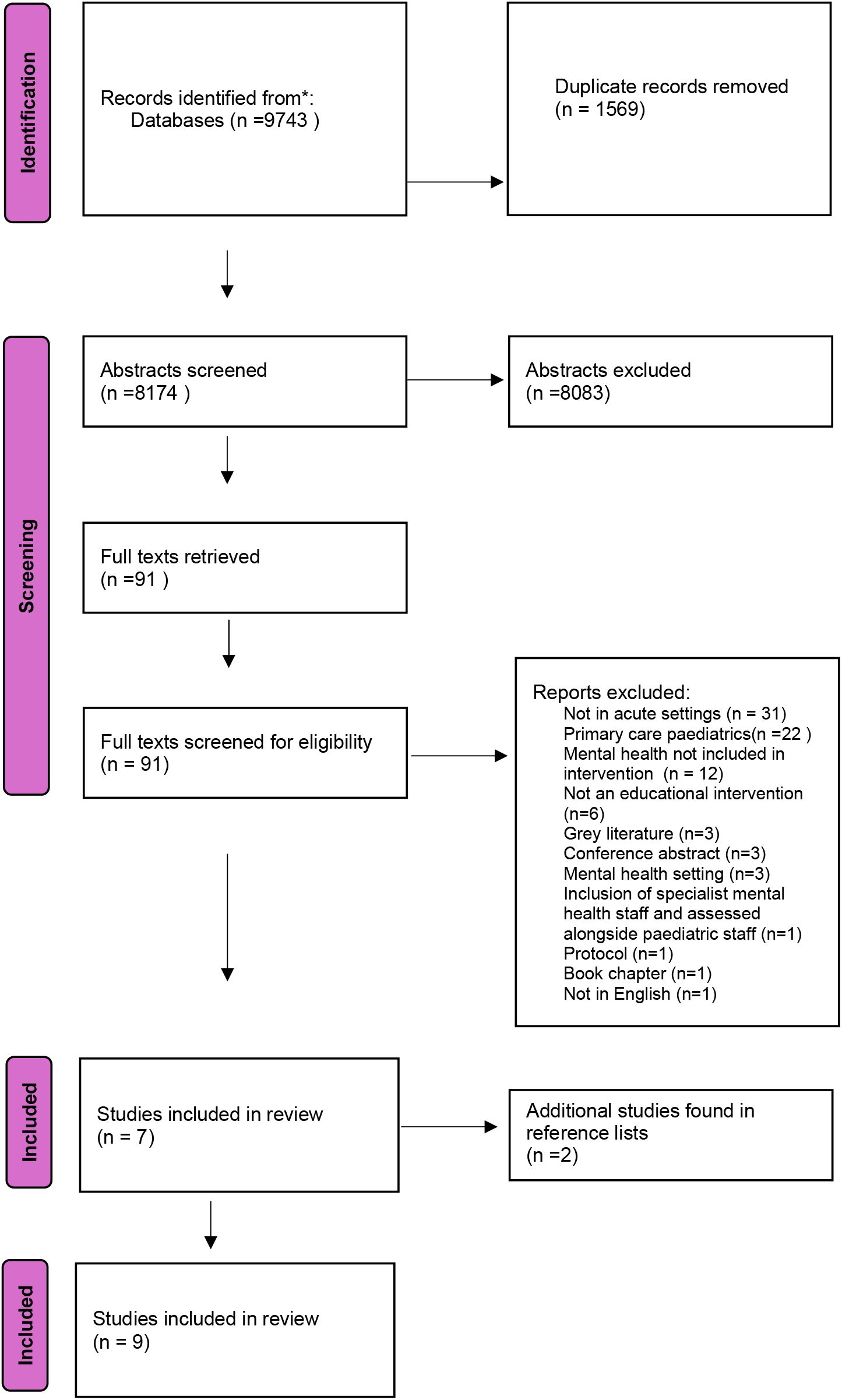
Prisma flow diagram

### Intervention Characteristics

Seven interventions were aimed at nursing staff,(31-36, 39) one focused on paediatric resident doctors,(37) and one on a wider mixed multidisciplinary team.(38)

Most interventions focused on improving professionals’ understanding and responsiveness to specific mental health presentations, namely challenging behaviour (4),(31, 33, 37, 38) self-harm (2),(34, 35) and anxiety and depression,(36) whereas one focussed on general mental health needs.(39) All interventions included content designed to improve staff attitudes towards mental health difficulties, most commonly via increasing confidence in assessment and management.(32–34, 36, 38) Some studies focused on improving communication (31) and risk assessment.(31, 38) One intervention was developed using social-cognitive theory as a framework model,(31) and another intervention utilised simulation-based education principles.(38) The timing of post intervention evaluations varied widely across studies, ranging from directly after (2),(31, 37) to 2 weeks(3),(33–35) 3-6 months (2) (36, 38) and 8 months after the intervention (1).(32)

Across studies, after classification, the BCT domains utilised were 2-9 (31–39) and 12.(31, 32) Interventions were evaluated using the following components of Kirkpatrick’s evaluation model included 2A (Learning-changes in attitudes) (8),(31-36, 38, 39) 2B (Learning-changes in knowledge/skills) (7),(31-33, 35-37, 39) and 3 (behavioural change) (3).(32, 35, 37)

### Quality Assessment

Quality assessments by study are summarised in Appendix II. First round inter-rater reliability between the two reviewers (AR and OS) was 92%, before final agreement for each study was made.

In the MMERSQI study design domain, most studies (31–36, 38) scored poorly with 9 or less (maximum possible score 23), most commonly for using only a single group pre-test and post-test design. While most studies scored maximum possible points for using appropriate statistical analysis according to the study design,(31, 32, 34–39) all studies scored poorly for complexity of statistical analysis (4 out of a maximum possible 8). Most frequently, this was owing to reliance on simple rather than inferential statistics.(31, 32, 34, 35, 37–39) In the majority of studies, evaluations were limited to ‘satisfaction, attitudes, perceptions, opinion and general facts’ assessments,(31, 33, 34, 36, 38) however 4 interventions used better measures of knowledge measured by low fidelity simulation or paper-based assessment.(32, 35, 37, 39)

### Summary of Interventions categorized by teaching techniques

#### Studies Applying Multimodal Intervention

Five studies used multiple teaching approaches in their intervention,(26, 31, 32, 36, 37, 39) including: didactic sessions,(31, 32, 36, 37, 39) skills sessions,(31, 32, 39) role play,(31, 36, 39) e-learning,(31, 32) observational learning via video demonstrations,(31, 37) Q&A sessions,(31, 36, 39) and interactive small group exercises.(37)

Mahoney et al(32) explored paediatric nurses’ self-reported confidence, knowledge, effectiveness, and use of strategies following training caring for autistic children with mental health needs. Participants received training on autism, communication strategies, sensory needs, and specific information resources delivered in multiple formats, including a 6-hour ‘train the trainer’ session. Confidence and effectiveness were measured using Likert scales ranging from 1 to 5(1= strongly disagree, 5= strongly agree).Post-intervention, participants reported feeling more confident using recommended resources with small-moderate effect sizes, (average 3.32 to 3.82, p<0.001, r=0.38. Participants felt more effective in delivering aspects of children’s care, including responding to challenging behaviour, regulating anxiety levels, and adapting to different sensory needs (small to moderate effect sizes in pre and post scores: average 3.45 to 3.69, p = 0.003, r=0.18).

Johnson et al(31) explored how staff knowledge and fear of working with children with developmental disabilities, predominantly autism, can be altered through an educational intervention for new nursing staff on preventing and managing challenging behaviours of this cohort. The intervention was developed using Bandura’s theory of self-efficacy, defined as “a person’s belief in his or her ability to perform a designated task”.(31) The 2-part intervention consisted of an online didactic session providing information on developmental disabilities in children, especially autism, suggesting strategies to prepare for procedures, communication and play, followed by video demonstrations, then opportunities to practice skills. After online education, participants rated their perceived knowledge and fear of caring for children with behaviour that challenges using a 10-point Likert scale. Participants reported increased knowledge relating to caring for children with challenging behaviours,(average pre-intervention 5.5, average post intervention 8.7), and decreased fear of managing children with challenging behaviours(pre 3.2, post 2.7).

Higson et al(36) developed an educational intervention for acute/general paediatric nurses to improve confidence of caring for children with eating disorders, self-harm and suicidal thoughts, anxiety and depression. This consisted of didactic lectures, role plays, testimonies from and a question and answer(Q&A) session with service users. Participants were asked to rate their confidence and knowledge/skills of managing children and young people presenting with mental health difficulties before and 3 months after the intervention using a 10-point Likert scale. Three months after the intervention, participants reported increased self-confidence in managing children and young people presenting with mental health difficulties(average pre 4.7, post 7.8), increased perceived knowledge of eating disorders(average pre 4.9, post 7.7), depression and anxiety(average pre 5.3, post 7.3), and self-harm(average pre 5.3 to 7.7).

Onilemo et al(39) tested an intervention to improve paediatric nurses’ attitudes and knowledge towards mental health using case control methodology. The 2-hour training included didactic lectures using the WHO mhGAP framework on child and adolescent mental health disorders.(40) This included teaching on communication skills and developing skills in psychoeducation through role play and a Q&A session. Before and after the intervention, participants completed an adapted version of the WHO knowledge and attitudes assessment of mental health disorders,(41) including questions on stigmatising attitudes. Each question was scored on a 3-point Likert scale (negative marking was applied with 0 for wrong answers, 1 for “I don’t know”, and 2 for correct answers. Participants demonstrated higher overall scores after completing the intervention, (average pre intervention 89.63 versus 95.36 post, p= 0.002), and higher post scores than controls (95.36 versus 86.14, p<0.001) with moderate effect sizes. There were no differences in the scores on questions assessing attitude following the or compared to controls.

Doja et al (37) explored the effects of the introduction of a mental health teaching programme for paediatric resident doctors’ knowledge of working with childhood disruptive behaviours. Following a needs assessment, four postgraduate paediatric training programmes taught paediatric resident doctors in assessment and management of disruptive behavioural disorders via didactic talks, video demonstrations of parent- and child-group therapy, and small group exercises. Change in knowledge was measured objectively by performance on multiple-choice questions(MCQ) pre and post intervention, and doctor behavioural change measured via an objective, structured clinical exam(OSCE) post intervention. Participant performance was compared to controls receiving education as usual. Post intervention scores on MCQ were better than pre and post (average 12.37 versus 16.82, p<0.001). There was no difference in OSCE scores between the cases and controls.

#### Studies Using Simulation

One study used simulation as its main teaching format.(38) Mitchell et al(38) explored whether healthcare professionals felt more confident to respond to aggressive behaviour in acute paediatric settings after participating in a simulation-based education programme. Participants had the opportunity to learn and practice using supportive communication techniques and de-escalation skills with a young person displaying signs of aggression on an inpatient ward. This was followed by a facilitated debrief for participants to analyse and reflect upon their actions. Participants self-rated their confidence responding to aggression pre-intervention, immediately post-intervention, and 3-6 months post-intervention using a 5-point Likert scale. Immediately post-intervention, newly qualified nurses on the graduate nurse program, felt more confident to manage clinical aggression, both for those who had worked with aggression before (average pre 2.25, 3.5, p<0.01), and those who had not(pre 2.19, post 3.63, difference p< 0.01).

#### E-learning

Two studies assessed a single e-learning programme in two different contexts (Our Care through Our Eyes).(34, 35) The intervention comprises of learning focusing on different aspects of caring for a young person presenting with self-harm through understanding self-harm, care pathways for CYP admitted to hospital, effective communication and assessing risk and managing safety. The program included didactic and interactive teaching.

Manning et al(35) studied the impact of the intervention on paediatric nurses’ knowledge, attitudes and behaviour towards self-harm. Information was gained on participant confidence using Likert scales, reporting the percentage of participants who stated they agreed or strongly agreed with particular statements. There were increases of proportions of participants in the domains of a) feeling more confident in caring for a CYP who has self-harmed increased post intervention (49% versus 78% p = 0.000), b) communicating with CYP and carers (64% vs 86% p=0.02), c) not making things worse for a CYP who has self-harmed (45% preintervention vs 68% postintervention, p =0.04). Although staff felt more confident in communicating effectively with a CYP who has self-harmed (56% vs 76%), this was not significant (p=0.06). Self-efficacy was measured using an adapted Self-Efficacy Towards Helping scale, a 10-item measure. Those who completed the intervention reported an unexplained non-significant decrease in self-efficacy (pre mean 28.81, post mean, p= 0.08), while total respondents demonstrated a significant decrease in self-efficacy (pre intervention mean 28.9, post intervention mean 27.6, p = 0.042, effect size 0.29). The authors did however find a significant increase in perceived effectiveness using a Likert scale (pre mean 9.88, post intervention mean 11.12, p= 0.008).

Singh-Weldon et al(34) also examined the ‘Our Care through Our Eyes’ for change in children’s nurses’ attitudes, beliefs, empathy, confidence and anxiety when caring for CYP who self-harm using a range of self-report questionnaires. After the intervention, non-significant positive changes in attitude were identified in participants, such as increased proportion of participants agreeing/strongly agreeing statement positive statements, such has “I help self-harming patients feel positive about themselves” (pre intervention 51%, post intervention 73%, statistical significant not tested for). Similarly there were non-significant positive findings regarding participant empathy, confidence and general attitude, alongside non-significant increases in feelings of anxiety.

#### Didactic teaching

A single study investigated the effects of an intervention delivered via didactic teaching.(33) Conley et al (33) developed an intervention to improve nurses’ knowledge and ease in caring for CYP with challenging behaviours. They used visual activity schedules, tools to be used for managing challenging behaviours, thought to provide structure and reduce anxiety for CYP.(33) The intervention was developed following a needs assessment. A greater proportion of participants reported increased comfort in caring for CYP with challenging behaviours using a Likert scale (average post intervention 60%, post 83%), increased perceived knowledge about diagnoses which increase risk for challenging behaviours (pre intervention 65%, post intervention 82%).

## DISCUSSION

To our knowledge, this is the first systematic synthesis of educational interventions for paediatric healthcare staff on acute general paediatric wards supporting CYP admitted with mental health needs. Overall, the quality of studies was low, and does not allow a single intervention to be recommended. However, valuable findings can be drawn from component elements of interventions, through findings related to the Kirkpatrick and BCT evaluation frameworks which may help select or design interventions to required changes for learner needs.

Using Kirkpatrick’s model for educational evaluation, we noted that interventions using certain teaching modalities may be more useful in leading to different outcomes. Successful change in attitudes (Kirkpatrick Level 2A) was reported in interventions using e-learning (35) and simulation.(38) Such attitudinal change could be particularly useful for learners where attitude change towards acute mental health is perceived as a desired outcome – and might be useful for example where there is staff ambivalence about role or pre-existing stigma.(1, 17) Successful change in knowledge and skills (Kirkpatrick level 2B) was reported as in three interventions using e-learning,(35) and two multimodal interventions,(37, 39) which included didactic talks, role play, question-and-answer sessions, and interactive small group exercises. These interventions may be useful for to generically increase overall knowledge and skills. Increases in perceived effectiveness in delivering care was associated with an e-learning based educational interventions. Some BCT approaches used in combination may be useful and more effective, for example, using the BCT 4.1 ‘instruction on how to perform a behaviour’ alongside BCT 6.1, ‘demonstration of behaviour’ appeared to positively impact emotions frequently described by staff when providing mental health care.(31, 33, 35) This included decreased fear(31) and increased confidence of caring for CYP with challenging behaviours,(33) and increased perceived effectiveness of caring for CYP following self-harm.(35)

A notable finding from our review was the absence of co-production with service users. This is an important omission as evidence suggests that co-production can ensure combining the realities of clinical practice with perspectives of those receiving care, challenge stigma and promote more compassionate and relational care.(42) Success has been reported in this approach for improving mental health care outside of the acute paediatric setting.(43, 44) Lack of co-production identified in our review may in part explain the quality of interventions, and highlights a need for future work to embed co-production within intervention design alongside evaluative frameworks such as Kirkpatrick and BCT.

Our review has several strengths. We used a priori inclusion and exclusion criteria with independent searches and quality assessments. Our research group team is multidisciplinary including professionals with relevant clinical, academic, educational and lived experience of mental illness and treatment in inpatient paediatric settings, enhancing its the epistemic and ecological validity by ensuring that interpretations of educational needs and intervention components were grounded in authentic patient perspectives and clinical realities. Our study also has limitations. Many studies were of small sample size, risking missing small effect sizes of interventions. A bigger limitation is that it is likely that we have not been able to include many existing interventions because anecdotally, interventions designed locally are often not written-up nor published, introducing publication bias. This could be tackled by prioritisation of faster local and national dissemination via conferences,(45) increasing numbers of academic and research active paediatricians,(46) and time in staff job plans to better assess and report newly introduced interventions.(47)

Our review allows for recommendations for future research, intervention development and reporting. We would recommend the use of a framework for reporting, ensuring interventions are adequately described, such as one developed by Meinema et al.(48) Most studies were conducted in specialist paediatric hospitals and settings, limiting applicability to more rural (“district”) general wards and calls for more research in these settings. Most studies focused primarily on nursing staff, and there is a need to develop more multidisciplinary interventions. Most interventions did not use an educational theory, implying they are likely to be based on “traditional and ritual”(20) rather than an established scientific framework. Some included studies were found to have used perception of learning (Kirkpatrick Level 2A) as an evaluation method which is problematic as there is an inherent discrepancy between subjective perceptions and objective measures of learning.(49, 50)

A major challenge in medical education is how to inspire the potentially ambivalent or reluctant learner who may not attend such interventions in the first place, other than making it mandatory. We found no particular interventions aimed to specifically target this issue, and it is possible that those who attended interventions were those motivated by the topic in the first place. As noted, some components of interventions using aspects of Kirkpatrick or BCT may induce behaviour and attitude change in part, but not necessarily get learners there in the first place. There are insights on this issue from a recent Cochrane review(51) which reported that interactive formats in educational interventions which focus on the benefits of improving serious or risky outcomes may be effective at improving attendance. Attendance and impact might also be simultaneously improved by utilising an ‘opinion leader’ or CYP perspectives, which aligns to the BCT technique of using a credible source in most of our included studies.(31–37, 39) Given the evidence that some professionals working on paediatric wards may not view their own role as relevant or necessary in supporting mental health admissions, exploring the role of professional identity in education, successfully applied in other contexts, may be potential factor in understanding needs, and improving interventions for overall care.(52, 53)

### Conclusion

Caring for patients admitted to general children’s wards for mental health concerns is important work with an important group of patients and an area that needs upskill. We present existing interventions, with components which may be valuable in improving care through education for specific targeting. There remains much to do to develop higher quality, framework based, and better evaluated studies utilising co-design with CYP and their carers, and also with the aim of getting a reluctant learner there in the first place.

## Data Availability

This is a systematic review therefore all extracted data is available in original papers, which are referenced.

## Data availability statement

All data relevant to the study are included in the article or uploaded as supplementary information.

## Ethics statements

Patient consent for publication: Not applicable

## Acknowledgements

We thank Veronica Phillips, Assistant Librarian at University of Cambridge Medical Library, for supporting this work.

## Contributors

LH, OS, AR and AG conceived and designed the review. OS and IP performed searches, and screened abstracts, titles and full texts with adjudication and support by LH and AR. OS and AR carried out the data extraction and quality assessment, with support from LH. MB and JRD provided clinical context, and JD and SP provided lived experience perspective and context. OS led the first draft, with LH and AR support. LH is PI of the grant and study from which the work originates. LO is part of the research team and led technology for the project. All authors contributed to the final draft.

---

| First author, year, country, setting, participants, type of mental health | Study design & aim of intervention | Length & format of intervention, length of follow-up | Facilitator, intervention focus, intervention content | Behaviour Change Techniques (BCTs) | Main Findings | Kirkpatrick's level of evaluation |
| --- | --- | --- | --- | --- | --- | --- |
| Conley et al., (2023), America<br>Children's hospital, 46 paediatric nurses, Children and Young People (CYP) at risk of challenging behaviours<br>Not co-designed | Mixed methods, Single arm pre-test post-test,<br>Increase knowledge of visual activity schedules (VAS) and nurse comfort in care of CYP at risk for challenging behaviours.<br>Developed following needs assessment with key stakeholders, which included nurses, child life specialists, and the clinical nurse specialist on an inpatient acute care unit in which many children with behavioural challenges related to ASD and other | Unknown<br>Written and/or in-person communication<br>2 weeks | Unknown<br>Knowledge & attitudes<br>The role of VAS for children at risk of challenging behaviours, benefits of use of VAS in hospital, and development of VAS in partnership with the family | 3.2 (social support-practical: nursing staff getting help from parents to make VAS), 4.1 (instruction on how to perform the behaviour: instruction on how to complete VAS), 5.1 (information about health consequences: information about the benefits of use of VAS in hospital setting), 5.3 (information about social and environmental consequences: VAS having the potential to increase understanding, facilitate participation, | Increased confidence in caring for children with challenging behaviours, knowledge of visual activity schedule, and of diagnoses of increasing risk of challenging behaviours. | Level 2A<br>Level 2B |
|  | diagnoses were admitted. Stakeholders agreed a knowledge gap existed regarding evidence-based tools to improve care provided to children with challenging behaviours. |  |  | support communication and increase independence, 6.1 (demonstration of the behaviour: project team provided demonstrations), 9.1 (credible source: signposted to the Child Life Specialist for further advice, delivered by nurse practitioners alongside medical student). |  |  |
| Doja et al., (2021), Canada<br>Unknown, 95 paediatric resident doctors, Childhood disruptive behavioural disorders<br>Not co- | Quantitative, Non-blinded cluster randomised trial, Increase knowledge in assessment and treatment of childhood disruptive behavioural disorders.<br>Curriculum developed by researchers, practitioners and knowledge users from | Length not stated Slides, video demonstrations, group exercises<br>Immediate | Paediatricians, child and adolescent psychiatrists<br>Knowledge Assessment of DBDs (disruptive behaviour disorders), psychosocial therapy recommendations, choice of | 4.1 (instruction on how to perform the behaviour: choice of pharmacological interventions), 5.1 information about health consequences: appropriate drug safety monitoring), 9.1 (credible source: delivered by paediatricians and child & adolescent | Significant improvement in knowledge (via test scores), no behavioural change (via OSCE) in group exposed to curriculum 6 months before the OSCE. | Level 2B<br>Level 3 |
| designed | across Canada. Needs assessment conducted among physicians in the assessment of disruptive behavioural disorders, psychosocial therapy recommendations, choice of pharmacological interventions, and appropriate drug safety monitoring. |  | pharmacological interventions, and appropriate drug monitoring. | psychiatrists) |  |  |
| Higson et al., (2016), UK Paediatric ward in hospital, 8 paediatric nurses, Anxiety, depression, self-harm and suicidal thoughts, eating disorders | Quantitative, Single arm pre-test post-test design, Improve knowledge and confidence of caring for CYP with MH problems.<br>Developed with external company 'Psychological Perspectives in Education and Primary Care' and adapted for frontline staff<br>Staff were previously asked what 3 topics they | 1 day PowerPoint presentation, role play, service user account and Q&A<br>3 months | Not specified<br>Knowledge, attitude-confidence<br>Improve knowledge and confidence of caring for CYP with MH problems | 4.1 (instructions on how to perform a behaviour, how to support young people with coping mechanisms for self-harm e.g. ice cubes), 5.1 (information about health consequences: self-harm & suicide, anxiety and depression and eating disorders), 8.1 behavioural practice: | Improved confidence and knowledge of eating disorders in all participants, most increased knowledge in depression, anxiety and self-harm. | Level 1<br>Level 2A<br>Level 2B |
| Not co-designed | would find useful as a starting point for teaching. |  |  | role play), 9.1 (credible source: experts by experience included) |  |  |
| Johnson et al., (2012), America, Children's hospital, 604 paediatric nurses and nursing allied professionals, Challenging behaviour<br>Not co-designed | Quantitative, Single arm pre-test post-test design, 1. Prevent and manage challenging behaviours via increased knowledge 2. decrease staff fear of caring for children with developmental disabilities. Developed by a team of security personnel, nurses, physicians, child life specialists and other healthcare professions, "most with personal and professional experience of working with children with developmental disabilities". Led by staff development advanced practice nurse.<br>No needs assessment | 2 hours (1-hour online learning, 1 hour instructor-led) Video demonstrations, Q&A, role play Immediate | Security personnel, child life specialists and nurses, Communication skills<br>1. Family centred care to prevent challenging behaviours<br>2. Communication strategies to prevent and manage challenging behaviours based on modified verbal judo | 2.2 (feedback on behaviour: practicing communication with colleagues and receive instructor feedback), 4.1(instruction on how to perform the behaviour: videos demonstrating techniques for distraction, communication, simple commands and management of over-stimulation), 6.1 (demonstration of the behaviour: videos with healthcare providers role modelling strategies for preventing or dealing with challenging | Improved knowledge of prevention and management of challenging behaviours, decreased fear | Level 2A<br>Level 2B |
|  | conducted |  |  | behaviours), 8.1<br>(behavioural practice<br>/rehearsal:<br>participants<br>practiced the<br>communication<br>methods with other<br>class participants<br>8.6 (generalisation of<br>target behaviour:<br>participants<br>practising strategies<br>in new situations 9.1<br>(credible source:<br>security personnel,<br>child life specialists,<br>and nurses, mostly<br>with<br>personal/professiona<br>l experience. Led by<br>advanced practice<br>nurse, 12.1<br>(restructuring the<br>physical<br>environment:<br>environmental<br>modifications<br>necessary to meet<br>the needs of children with developmental disabilities at risk for challenging behaviours, including minimising lights and noise, decreasing clutter and odours) |  |  |
| Mahoney et al., (2023), America Children's hospital, 300 paediatric nurses and nursing aides, Autism and mental health | Quantitative, Single arm pre-test post-test design. Increase knowledge, efficacy and confidence of nursing staff to improve MH of children with ASD.<br>Developed by hospital administrators with clinical backgrounds (occupational therapy practitioners, speech-language pathologists, physicians, nurses, child life specialists and behavioural therapists). Needs assessment | 15 minutes-6 hours. Four options: 6-hour train the trainer, 1-hour overview presentation, 15-minute orientation (led by nurses who had completed the 6-hour training), online training, Up to 8 months | OT practitioners, Speech and Language pathologists, physicians nurses, behavioural therapists child life specialists, Knowledge, attitudes (confidence), communication skills. Information about ASD, communication strategies, sensory needs | 4.1 (instruction on how to perform the behaviour: communication strategies), 5.1 (information about health consequences: information about ASD), 5.3 (information about social and environmental consequences: sensory needs and strategies) 7.5 (remove aversive stimulus: environmental | Significant improvement in self-efficacy, effectiveness, confidence and strategies used generally and in paired sample. Changes in knowledge not significant | Level 2A<br>Level 2B<br>Level 3 |
|  | completed in collaboration with a university partner. |  | and strategies, Adaptive Care program criteria, resources and referral process. 1 case example in all except 15-minute training | modifications), 9.1 (credible source: occupational therapy practitioners, speech-language pathologists, physicians, nurses, child life specialists and behavioural therapists), 12.3 (Avoidance /reducing exposure to cues for the behaviour: information gathered about what triggered negative reaction in child). |  |  |
| Manning et al., (2017), UK Children's hospital, 98 paediatric nurses<br>Self harm, risk assessment | Mixed methods<br>Single arm pre-test post-test design,<br>Impact on knowledge, attitudes confidence and behavioural intention of working with CYP with self-harm.<br>No specific needs | 45 minutes<br>E-learning, activities, self-assessment<br>Up to 2 weeks | Self-directed e-learning, Knowledge, communication, application. Understanding self-harm and care pathways, effective communication | 4.1 (instruction on how to perform the behaviour: video demonstrations including assessing risk and managing safety with CYP admitted with self-harm), 6.1 (demonstration of | Increased effectiveness and knowledge of self-harm in total responders and intervention completers. Higher effect size in intervention completers vs total | Level 2A<br>Level 2B<br>Level 3 |
| Co-designed by children and young people, registered children's nurses and academics | assessment, based on previous research which shows children's nurses lack communication skills with CYP with MH problems. |  | with CYP following self-harm admission, Assessing risk and managing safety | the behaviour; video demonstrations including effective communication with CYP following self-harm admission), 9.1 (credible source: co-produced with CYP service users, registered children's nurses and academics) | responders |  |
| Mitchell et al., (2020), Australia<br>Children's hospital, 146 hospital paediatric staff, Clinical aggression<br>Not co-designed | Mixed methods, Single arm pre-test post-test design, Perceived confidence in managing clinical aggression. Simulations developed by study investigators (experience not stated) in conjunction with simulation faculty. No needs assessment. | 2 hours, Simulation and debrief, 3-6 months | Facilitated by simulation faculty member (background unknown)<br>Not specified, Attitude-confidence, communication, de-escalation. Communication, de-escalation skills | 2.2 (feedback on behaviour: via debrief), 4.1 (instruction on how to perform the behaviour: communication strategies for managing aggression), 6.2 (social comparison: watching others performing roles in simulation exercise), 8.1 (behavioural practice/rehearsal: | Significant improvement in confidence managing clinical aggression | Level 1<br>Level 2A |
|  |  |  |  | scenario enabled participants to apply supportive communication techniques and de-escalation skills), 8.6 (generalisation of target behaviour; participants repeated skills in two different scenarios) |  |  |
| Onilemo et al., (2021),<br>Nigeria<br>Paediatric ward, teaching hospital,<br>156 paediatric nurses,<br>General mental health<br>Not co-designed | Quantitative,<br>Two-group pretest-post test trial, block randomisation,<br>Determine the effect of a mental health training intervention on the knowledge and attitudes of nurses work on acute paediatric wards.<br>Intervention taken from the WHO mhgap general framework on child and adolescent mental health disorders and the WHO mhgap behavioural disorders module. No | 2 hours<br>Didactics, role play, Q&A<br>Not reported | Child and adolescent psychiatrist, mental health nurse,<br>Knowledge, communication skills, application.<br>Training on recognition, aetiology and management of common MH disorders, communication with children and families, how to | 4.1 (instruction on how to perform the behaviour: strong emphasis on communicating with children and their families, and how to carry out psychoeducation), 5.1 (information about health consequences: taught about the presentations of disorders), 8.1 (behavioural | Significantly increased knowledge in intervention group compared to control and compared to baseline score in intervention group. No change in attitude between or within groups | Level 2A<br>Level 2B |
|  | needs assessment |  | carry out psychoeducation | practice/rehearsal: information given about psychoeducation and role plays), 9.1 (credible source: taught by psychiatrist and mental health nurse) |  |  |
| Singh-Weldon et al., (2022), UK Children's hospital, 42 paediatric nurses, Self-harm<br>Co-designed, but not developed by study group | Mixed methods, Single arm pre-test post-test design.<br>Designed by children and young people, registered children's nurses and academics | 45 minutes, E-learning, interactive exercises<br>Up to 2 weeks | Self-directed, Attitude-confidence, anxiety, empathy. Influence on children's nurses' attitudes, beliefs, empathy, confidence and anxiety in relation to the care of CYP who self-harm. | 4.1 (instruction on how to perform the behaviour: video demonstrations including assessing risk and managing safety with CYP admitted with self-harm), 6.1 (demonstration of the behaviour; video demonstrations including effective communication with CYP following self-harm admission), 9.1 (credible source: co-produced with CYP service users,<br>registered children's<br>nurses and<br>academics) | Non-significant changes in attitudes, empathy, anxiety and confidence | Level 2A |

## References

1. Vázquez-Vázquez A, Smith A, Gibson F, Roberts H, Mathews G, Ward JL, et al. Admissions to paediatric medical wards with a primary mental health diagnosis: a systematic review of the literature. Arch Dis Child. 2024;109(9):707–16.

2. Gallagher KAS, Bujoreanu IS, Cheung P, Choi C, Golden S, Brodziak K, et al. Psychiatric Boarding in the Pediatric Inpatient Medical Setting: A Retrospective Analysis. Hosp Pediatr. 2017;7(8):444–50.

3. Ward JL, Vázquez-Vázquez A, Phillips K, Settle K, Pilvar H, Cornaglia F, et al. Admission to acute medical wards for mental health concerns among children and young people in England from 2012 to 2022: a cohort study. Lancet Child Adolesc Health. 2025;9(2):112–20.

4. Clisu DA, Layther I, Dover D, Viner RM, Read T, Cheesman D, et al. Alternatives to mental health admissions for children and adolescents experiencing mental health crises: A systematic review of the literature. Clin Child Psychol Psychiatry. 2022;27(1):35–60.

5. Otis M, Barber S, Amet M, Nicholls D. Models of integrated care for young people experiencing medical emergencies related to mental illness: a realist systematic review. Eur Child Adolesc Psychiatry. 2023;32(12):2439–52.

6. Hssib. Investigation Report Keeping children and young people with mental health needs safe: the design of the paediatric ward 2024.

7. Manning JC, Carter T, Walker G, Coad J, Aubeeluck A. Assessing risk of self-harm in acute paediatric settings: a multicentre exploratory evaluation of the CYP-MH SAPhE instrument. BMJ open. 2021;11(5):e043762.

8. Mitchell M, Newall F, Williams K. Behavioural emergencies in a paediatric hospital environment. J Paediatr Child Health. 2022;58(6):1033–8.

9. Chapman S, Hudson LD, Street KN. Restrictive eating disorders in children and young people: the role of the paediatrician and paediatric ward. Arch Dis Child. 2023;108(4):e4.

10. Hawley M, O’Neill J, Dorland J, Richards S, Kinney S, Court A, Rayner C. Restraint for nasogastric tube feeding in young people with anorexia nervosa or atypical anorexia nervosa: a retrospective audit. J Eat Disord. 2025;13(1):143.

11. Hampton E, Richardson JE, Bostwick S, Ward MJ, Green C. The current and ideal state of mental health training: pediatric resident perspectives. Teach Learn Med. 2015;27(2):147–54.

12. Buckley S. Caring for those with mental health conditions on a children’s ward. Br J Nurs. 2010;19(19):1226–30.

13. Wu WL, Chen SL. Nurses’ perceptions on and experiences in conflict situations when caring for adolescents with anorexia nervosa: A qualitative study. Int J Ment Health Nurs. 2021;30 Suppl 1:1386–94.

14. Lakeman R, McIntosh C. Perceived confidence, competence and training in evidence-based treatments for eating disorders: a survey of clinicians in an Australian regional health service. Australas Psychiatry. 2018;26(4):432–6.

15. Hudson LD, Chapman S, Street KN, Nicholls D, Roland D, Dubicka B, et al. Increased admissions to paediatric wards with a primary mental health diagnosis: results of a survey of a network of eating disorder paediatricians in England. Arch Dis Child. 107. England 2022. p. 309–10.

16. Watson E. CAMHS liaison: supporting care in general paediatric settings. Paediatr Nurs. 2006;18(1):30–3.

17. Sukhera J, Miller K, Milne A, Scerbo C, Lim R, Cooper A, Watling C. Labelling of mental illness in a paediatric emergency department and its implications for stigma reduction education. Perspect Med Educ. 2017;6(3):165–72.

18. Worsley D, Barrios E, Shuter M, Pettit AR, Doupnik SK. Adolescents’ Experiences During “Boarding” Hospitalization While Awaiting Inpatient Psychiatric Treatment Following Suicidal Ideation or Suicide Attempt. Hosp Pediatr. 2019;9(11):827–33.

19. Wilkes M, Bligh J. Evaluating educational interventions. Bmj. 1999;318(7193):1269–72.

20. Artino AR, Jr., Konopasky A. The Practical Value of Educational Theory for Learning and Teaching in Graduate Medical Education. J Grad Med Educ. 2018;10(6):609–13.

21. Kirkman MA, Sevdalis N, Arora S, Baker P, Vincent C, Ahmed M. The outcomes of recent patient safety education interventions for trainee physicians and medical students: a systematic review. BMJ Open. 2015;5(5):e007705.

22. Kulju E, Jarva E, Oikarinen A, Hammarén M, Kanste O, Mikkonen K. Educational interventions and their effects on healthcare professionals’ digital competence development: A systematic review. Int J Med Inform. 2024;185:105396.

23. Menezes P, Guraya SY, Guraya SS. A Systematic Review of Educational Interventions and Their Impact on Empathy and Compassion of Undergraduate Medical Students. Front Med (Lausanne). 2021;8:758377.

24. Hill J, Gratton N, Kulkarni A, Hamer O, Harrison J, Harris C, et al. The effectiveness of evidence-based healthcare educational interventions on healthcare professionals’ knowledge skills, attitudes, professional practice and healthcare outcomes: Systematic review and meta-analysis. J Eval Clin Pract. 2024;30(6):909–35.

25. Page M, McKenzie, J., Bossuyt, P., Boutron, I., Hoffman, T., Mulrow, C., Shamseer, L., Tetzlaff, J.,. The PRISMA 2020 statement: an updated guideline for reporting systematic reviews. BMJ. 2021.

26. Al Asmri M, Haque MS, Parle J. A Modified Medical Education Research Study Quality Instrument (MMERSQI) developed by Delphi consensus. BMC Med Educ. 2023;23(1):63.

27. Lawson McLean A, Schwarz F. Quantifying the Quality of Medical Education Studies: The MMERSQI Approach. BMC Medical Education. 2023;23(1):320.

28. Michie S, Richardson M, Johnston M, Abraham C, Francis J, Hardeman W, et al. The behavior change technique taxonomy (v1) of 93 hierarchically clustered techniques: building an international consensus for the reporting of behavior change interventions. Ann Behav Med. 2013;46(1):81–95.

29. Smidt A, Balandin S, Sigafoos J, Reed VA. The Kirkpatrick model: A useful tool for evaluating training outcomes. J Intellect Dev Disabil. 2009;34(3):266–74.

30. Kirkpatrick DL. The Four Levels of Evaluation. In: Brown SM, Seidner CJ, editors. Evaluating Corporate Training: Models and Issues. Dordrecht: Springer Netherlands; 1998. p. 95–112.

31. Johnson NL, Lashley J, Stonek AV, Bonjour A. Children with developmental disabilities at a pediatric hospital: staff education to prevent and manage challenging behaviors. J Pediatr Nurs. 2012;27(6):742–9.

32. Mahoney WJ, Abraham G, Villacrusis M. Many Hands Working Together: Adapting Hospital Care to Support Autistic Children’s Mental Health. Am J Occup Ther. 2023;77(2).

33. Conley CR, Wendt L, Schindler CA. Improving Nurses’ Knowledge in Caring for Children with Challenging Behaviors. Pediatric Nursing. 2023;49(3):142–7.

34. Singh-Weldon J, Tsianakas V, Murrells T, Grealish A. Preparing children’s nurses for working with children and adolescents who self-harm: Evaluating the ‘our care through our eyes’ e-learning training package. Int J Ment Health Nurs. 2022;31(6):1427–37.

35. Manning JC, Carter T, Latif A, Horsley A, Cooper J, Armstrong M, et al. ‘Our Care through Our Eyes’. Impact of a co-produced digital educational programme on nurses’ knowledge confidence and attitudes in providing care for children and young people who have self-harmed: a mixed-methods study in the UK. BMJ Open. 2017;7(4):e014750.

36. Higson J, Emery A, Jenkins M. Improving children’s nurses’ knowledge of caring for people with mental health problems. Nurs Child Young People. 2017;29(1):25–9.

37. Doja A, Pringsheim T, Andrade BF, Cowley L, Healy SA, Baron T, et al. Implementation and evaluation of a curriculum on the assessment and treatment of disruptive behaviour disorders. Paediatr Child Health. 2021;26(8):458–61.

38. Mitchell M, Newall F, Sokol J, Heywood M, Williams K. Simulation-based education to promote confidence in managing clinical aggression at a paediatric hospital. Adv Simul (Lond). 2020;5:21.

39. Onileimo V, Bella-Awusah T, Lasebikan V, Omigbodun O. Brief training in child and adolescent mental health: Impact on the knowledge and attitudes of pediatric nurses in Nigeria. J Child Adolesc Psychiatr Nurs. 2021;34(3):164–70.

40. Team WHO. mhGAP Intervention Guide Version 2.0 for mental, neurological and substance use disorders in non-specialized health settings 2016.

41. Bella-Awusah TAB, Dogra N, Omigbodun O. The impact of a mental health teaching programme on rural and urban secondary school students’ perceptions of mental illness in southwest Nigeria. J Child Adolesc Ment Health. 2014;26(3):207–15.

42. Downs J. Beyond the methodological binary: coproduction as the third pillar of mental health science BMJ Mental Health 2025;28(1).

43. Horgan A, Manning, F., Bocking, J., Happell, B., Lahti, M., Doody, R., & Biering, P. ‘To be treated as a human’: Using co-production to explore experts by experience involvement in mental health nursing education The COMMUNE project. International journal of mental health nursing. 2018;27(4):1282–91.

44. Norton MJ, & Swords, C. Creating equality for those in crisis: transforming acute inpatient mental health services through co-production Academic Quarter| Akademisk kvarter. 2021;23:64–79.

45. Van Cleave J, Dougherty D, Perrin JM. Strategies for addressing barriers to publishing pediatric quality improvement research. Pediatrics. 2011;128(3):e678–86.

46. Mustafa K, Murray CC, Nicklin E, Glaser A, Andrews J. Understanding barriers for research involvement among paediatric trainees: a mixed methods study. BMC Med Educ. 2018;18(1):165.

47. Torres C, Mendes F, Duarte AM, Vilaça S, Barbieri-Figueiredo MdC. Implementation of evidence-based practice in paediatric nursing care: Facilitators and barriers. Collegian. 2024;31(5):342–7.

48. Meinema JG BN, van Etten-Jamaludin FS, Visser MRM, van Dijk N. Intervention Descriptions in Medical Education: What Can Be Improved? A Systematic Review and Checklist. Acad Med. 2019;94(2):281–90.

49. Barnsley L, Lyon PM, Ralston SJ, Hibbert EJ, Cunningham I, Gordon FC, Field MJ. Clinical skills in junior medical officers: a comparison of self-reported confidence and observed competence. Med Educ. 2004;38(4):358–67.

50. Persky AM, Lee E, Schlesselman LS. Perception of Learning Versus Performance as Outcome Measures of Educational Research. Am J Pharm Educ. 2020;84(7):ajpe7782.

51. Forsetlund L, O’Brien MA, Forsén L, Reinar LM, Okwen MP, Horsley T, Rose CJ. Continuing education meetings and workshops: effects on professional practice and healthcare outcomes. Cochrane Database Syst Rev. 2021;9(9):Cd003030.

52. Pach J, Stoffels, M., Schoonmade, L., van Ingen, E., Kusurkar, R.,. The impact of educational activities on professional identity formation in social sciences and humanities: a scoping review. Educational Research Review. 2025;48.

53. Wald H. Professional identity (trans)formation in medical education: reflection, relationship, resilience. Acad Med. 2015;90(6):701–6.

