## Appendix 1 and 2 for "What works in educational interventions for teams caring for children and young people admitted to general acute paediatric wards for mental health? A systematic review"

SUPPLEMENTARY DATA

**Appendix I: Database Search Terms**

***PubMed:***

Search #1: (((((child*[Title/Abstract]) OR (adolescen*[Title/Abstract])) OR (youth[Title/Abstract])) OR ("young person"[Title/Abstract])) OR ("young people"[Title/Abstract])) OR (teen*[Title/Abstract])

Search #2: (paedia*[Title/Abstract]) OR (pedia*[Title/Abstract])

Search #3: (((((((training[Title/Abstract]) OR (educat*[Title/Abstract])) OR (workshop*[Title/Abstract])) OR (skills[Title/Abstract])) OR (class*[Title/Abstract])) OR (learn*[Title/Abstract])) OR (teach*[Title/Abstract])) OR (taught[Title/Abstract])

Search #4: ((((((doctor*[Title/Abstract]) OR (residen*[Title/Abstract])) OR (nurs*[Title/Abstract])) OR ("healthcare assistan*"[Title/Abstract])) OR ("healthcare professional*"[Title/Abstract])) OR (physio*[Title/Abstract])) OR (therap*[Title/Abstract])

Search #5: deleted search

Search #6: ((((((((((((((((("mental health"[Title/Abstract]) OR (psych*[Title/Abstract])) OR ("self harm"[Title/Abstract])) OR (anorex*[Title/Abstract])) OR ("eating disorder"[Title/Abstract])) OR (agitat*[Title/Abstract])) OR (dysregulat*[Title/Abstract])) OR ("emotional dysregulat*"[Title/Abstract])) OR (behavio*[Title/Abstract])) OR ("challenging behavio*"[Title/Abstract])) OR ("disrupt* behav*"[Title/Abstract])) OR (autis*[Title/Abstract])) OR (psychosis[Title/Abstract])) OR (tic[Title/Abstract])) OR (depress*[Title/Abstract])) OR (anxi*[Title/Abstract])) OR ("substance abuse"[Title/Abstract])) OR ("substance misuse"[Title/Abstract])

Search #7: 2 and 4

(((((((doctor*[Title/Abstract]) OR (residen*[Title/Abstract])) OR (nurs*[Title/Abstract])) OR ("healthcare assistan*"[Title/Abstract])) OR ("healthcare professional*"[Title/Abstract])) OR (physio*[Title/Abstract])) OR (therap*[Title/Abstract])) AND ((paedia*[Title/Abstract]) OR (pedia*[Title/Abstract]))

Search #8:7 and 3 and 6 and 1

((((((((((doctor*[Title/Abstract]) OR (residen*[Title/Abstract])) OR (nurs*[Title/Abstract])) OR ("healthcare assistan*"[Title/Abstract])) OR ("healthcare professional*"[Title/Abstract])) OR (physio*[Title/Abstract])) OR (therap*[Title/Abstract])) AND ((paedia*[Title/Abstract]) OR (pedia*[Title/Abstract]))) AND ((((((((training[Title/Abstract]) OR (educat*[Title/Abstract])) OR (workshop*[Title/Abstract])) OR (skills[Title/Abstract])) OR (class*[Title/Abstract])) OR (learn*[Title/Abstract])) OR (teach*[Title/Abstract])) OR (taught[Title/Abstract]))) AND (((((((((((((((((("mental health"[Title/Abstract]) OR

(psych*[Title/Abstract])) OR ("self harm"[Title/Abstract])) OR (anorex*[Title/Abstract])) OR ("eating disorder"[Title/Abstract])) OR (agitat*[Title/Abstract])) OR (dysregulat*[Title/Abstract])) OR ("emotional dysregulat*"[Title/Abstract])) OR (behavio*[Title/Abstract])) OR ("challenging behavio*"[Title/Abstract])) OR ("disrupt* behav*"[Title/Abstract])) OR (autis*[Title/Abstract])) OR (psychosis[Title/Abstract])) OR (tic[Title/Abstract])) OR (depress*[Title/Abstract])) OR (anxi*[Title/Abstract])) OR ("substance abuse"[Title/Abstract])) OR ("substance misuse"[Title/Abstract]))) AND ((((((child*[Title/Abstract]) OR (adolescen*[Title/Abstract])) OR (youth[Title/Abstract])) OR ("young person"[Title/Abstract])) OR ("young people"[Title/Abstract])) OR (teen*[Title/Abstract])

*MeSH Terms for PubMed*

Search 2: Pediatrics

Search 3: (education, medical), (education, nursing), learning health system

Search 4: pediatricians, nurses, physical therapists , health personnel

Search 6: child mental disorder, tics, depression, substance-related disorders, self-injurious behavior

***ERIC:***

((Doctor OR resident OR nurse OR healthcare assistant OR healthcare professional OR physio OR therapist) AND (paediatric OR pediatric OR child OR adolescent))

AND

((training OR education OR workshop OR skills OR class OR learn OR teach OR taught)

AND

("mental health" OR psych OR "self harm" OR anorexia OR "eating disorder" OR agitation OR dysregulation OR "emotional dysregulation" OR behaviour OR behavior OR "challenging behaviour" OR "challenging behavior" OR "disruptive behaviour" OR "disruptive behavior" OR autism OR psychosis OR tic OR depression OR anxiety OR "substance abuse" OR "substance misuse") AND (child OR adolescen OR youth OR "young person" OR "young people" OR teen))

***Web of Science:***

SEARCH 1: child* OR adolescen* OR youth OR “young person” OR “young people” OR teen*

SEARCH 2: paedia* OR pedia*

SEARCH 3: training OR educat* OR workshop* OR skills OR class* OR learn* OR teach* OR taught

SEARCH 4: doctor* OR residen* OR nurs* OR “healthcare assistan*” OR “healthcare professional” OR physio* OR therap*

SEARCH 5: “mental health” OR psych* OR “self-harm” OR anorex* OR “eating disorder” OR agitat* OR dysregulat* OR “emotional dysregulat*” OR behavio* OR “challenging behavio*” OR disrupt* OR autis* OR psychosis OR tic OR depress* OR anxi* OR “substance abuse” OR “substance misuse”

SEARCH 6: #2 AND #4

SEARCH 7: #1 AND #3 AND #6 AND #5

***PsychInfo***

Search 1: child*.mp.

Search 2: adolescen*.mp.

Search 3: youth.mp.

Search 4: “young person”.mp.

Search 5: “young people”.mp.

Search 6: teen*.mp.

Search 7: or/1-6

Search 8: paedia*.ti,ab,id,mf.

Search 9: pedia*.ti,ab,id,mf.

Search 10: or/8-9

Search 11: training.ti,ab,id,mf

Search 12: educat*.ti,ab,id,mf

Search 13: workshop*.ti,ab,id,mf

Search 14: skills.ti,ab,id,mf

Search 15: class*.ti,ab,id,mf

Search 16: learn*.ti,ab,id,mf

Search 17: teach*.ti,ab,id,mf

Search 18: taught.ti,ab,id,mf

Seach 19: or/11-18

Search 20: doctor*.ti,ab,id,mf

Search 21: residen*.ti,ab,id,mf

Search 22: nurs*.ti,ab,id,mf

Search 23: “healthcare assistan*.ti,ab,id,mf

Search 24: “healthcare professional” .ti,ab,id,mf

Search 25: physio*.ti,ab,id,mf

Search 26: therap*.ti,ab,id,mf

Search 27: or/20-26

Search 28: “mental health” .ti,ab,id,mf

Search 29: psych*.ti,ab,id,mf

Search 30: “self harm” .ti,ab,id,mf

Search 31: anorex*.ti,ab,id,mf

Search 32: “eating disorder” .ti,ab,id,mf

Search 33: agitat*.ti,ab,id,mf

Search 34: dysregulat*.ti,ab,id,mf

Search 35: “emotional dysregulat*”.ti,ab,id,mf

Search 36: behavio*.ti,ab,id,mf

Search 37: “challenging behavio*”.ti,ab,id,mf

Search 38: “disrupt behav*”.ti,ab,id,mf

Search 39: autis*.ti,ab,id,mf

Search 40: psychosis.ti,ab,id,mf

Search 41: tic.ti,ab,id,mf

Search 42: depress*.ti,ab,id,mf

Search 43: anxi*.ti,ab,id,mf

Search 44: “substance abuse” .ti,ab,id,mf

Search 45: “substance misuse” .ti,ab,id,mf

Search 46: or/28-45

Search 47: 10 and 27

Search 48: 7 and 19 and 46 and 47

Search 49: 47 and 19 and 46 and 7

**Appendix II: Quality Assessment**

| **MMERSQI Item** | Conley  et al.,  2023 | Doja et al.,  2021 | Higson et al.,  2016 | Johnson et al.,  2012 | Mahoney et al.,  2023 | Manning et al., 2017 | Mitchell  et al.,  2020 | Onilemo et al.,  2021 | Singh-Weldon et al.,  2022 |
| --- | --- | --- | --- | --- | --- | --- | --- | --- | --- |
| 1. Study design | 9 | 23 | 9 | 9 | 9 | 9 | 9 | 16 | 9 |
| 2. Sampling – power calculation | 0 | 0 | 0 | 0 | 0 | 0 | 0 | 3 | 3 |
| 3. Sampling – participant characteristics | 0 | 0 | 0 | 0 | 3 | 3 | 3 | 3 | 3 |
| 4. Sampling – response rate | 1 | 1 | 4 | 2 | 1 | 1 | 4 | 4 | 2 |
| 5. Setting | 5 | 5 | 5 | 5 | 5 | 5 | 5 | 5 | 5 |
| 6. Type of data | 4 | 11 | 4 | 4 | 6 | 4 | 4 | 6 | 4 |
| 7. Validity of evaluation instrument-internal  structure | 0 | 0 | 0 | 0 | 0 | 5 | 0 | 0 | 5 |
| 8. Validity of evaluation instrument-content | 0 | 5 | 0 | 0 | 5 | 5 | 0 | 5 | 5 |
| 9. Validity of evaluation instrument – relation to other variables | 0 | 0 | 0 | 0 | 0 | 0 | 0 | 0 | 0 |
| 10. Appropriateness of the analysis | 0 | 9 | 9 | 9 | 9 | 9 | 9 | 9 | 9 |
| 11. Complexity of the analysis | 4 | 4 | 4 | 4 | 4 | 4 | 4 | 4 | 4 |
| 12. Outcomes | 7 | 9 | 7 | 7 | 8 | 9 | 7 | 9 | 7 |
| **TOTAL** | **30** | **67** | **42** | **40** | **50** | **54** | **45** | **64** | **56** |

*Table 2: Quality Assessment using Modified MMERSQI Tool*

MMERSQI scoring criteria:

**Study design**

a. Single group cross-sectional or single group post-test only = 7

b. Single group pre-test and post-test = 9

c. Nonrandomised, 2 groups = 10

d. Randomised controlled trial with high-risk bias = 11

e. Randomised controlled trial with moderate risk bias = 16

f. Randomised controlled trial with low-risk bias = 23

**Sampling**

2. Is there a power calculation (sufficient statistical power) for sample size?

a. No = 0

b. Yes=	3

3. Are detailed participant characteristics for each arm reported?

a. No =	0

b. Yes =	3

4. Response rate, % (select one)

a. Not reported = 0.5

b. <50 = 1

c. 50-74 = 2

d. >75 = 4

**Setting**

5. Institutions studied: (please select one)

a. Single Centre = 5

b. Multicentre, no further specification = 5

c. Multicentre with specification but not appropriate/balanced/complementary = 5

d. Multicentre with appropriate and balanced/complementary = 8

**Type of Data**

6. Type of Data : (please select one)

Assessment by participants (self-assessment) = 4

Objective measurements:

a. Knowledge test (e.g. recall type questions) = 6

b. Applied knowledge test (e.g. analysis and problem-solving type questions) = 8

c. Skills = 11

**Validity of Evaluation Instrument**

7. Internal structure

a. Not applicable

b. Not reported = 0

c. Reported= 5

8. Content

a. Not applicable

b. Not reported = 0

c. Reported = 5

9. Relationship to other variables

a. Not applicable

b. Not reported = 0

a. Reported = 5

**Data analysis**

10. Appropriateness of analysis

a. Inappropriate for study design or type of data = 0

b. Appropriate for study design, type of data =	9

11. Complexity of analysis (if appropriate for study design), please select one:

a. Descriptive analysis only = 4

b. Simple inferential statistics = 4

c. Modelling and more complex analysis = 8

**Outcomes**

12. Satisfaction, attitudes, perceptions, opinions, general facts = 7

Knowledge, measured by (please select one):

a. Low fidelity simulation or paper-based assessments = 9

b. High fidelity simulation = 12

Skills measured by: (please choose one)

a. Low fidelity simulation or paper-based assessments = 8

b. High fidelity simulation = 12

Behaviours in clinical environment =13

Patient/health outcome =
